# Social Determinants of Sepsis Mortality in the United States: A Retrospective, Epidemiologic Analysis

**DOI:** 10.1101/2024.12.19.24319343

**Authors:** Ahad Khaleghi Ardabili, Alireza Vafaei Sadr, Vida Abedi, Anthony S Bonavia

**Author notes:** Corresponding author: Anthony Bonavia, MD FCCP.

## Abstract

**Objective:** To determine whether neighborhood-level social determinants of health (SDoH) influence mortality following sepsis in the United States.

**Study Setting and Design:** Retrospective analysis of data from 4.4 million hospitalized patients diagnosed with sepsis, identified using International Classification of Diseases-10 codes, across the United States.

**Data Sources and Analytic Sample:** De-identified, aggregated data were sourced from the TriNetX Diamond Network. SDoH variables included income, housing cost burden, broadband access, park proximity, racial/ethnic diversity, and the Area Deprivation Index (ADI). The primary outcome was mortality, assessed using univariate and multivariate binomial generalized linear models. Predictors with high multicollinearity (Variance Inflation Factor > 5) were excluded to enhance model stability.

**Principal Findings:** Lower median income, higher ADI scores, limited park access, and lack of broadband connectivity were strongly associated with increased sepsis mortality. Unexpectedly, greater racial/ethnic diversity was negatively associated with mortality, possibly reflecting regional disparities in healthcare access and socioeconomic conditions. Multivariate analyses revealed that the inclusion of SDoH variables attenuated some effects observed in univariate models, highlighting their complex interplay. Random Forest analysis identified park access as the most important predictor of sepsis mortality, emphasizing its role as a potential proxy for broader neighborhood resources.

**Conclusions:** Neighborhood-level SDoH are critical for risk stratification in sepsis prognostic models and should be systematically integrated into predictive frameworks. These findings highlight the need for targeted public health interventions to address social vulnerabilities, enhance access to green spaces, and reduce disparities in sepsis outcomes across diverse populations.

**Callout Box:** *What is known on this topic:* Social determinants of health (SDoH) influence a variety of health outcomes although their impact on sepsis mortality has been under-explored. Sepsis is a leading cause of mortality in the United States, with over 350,000 deaths annually.

*What this study adds:* Neighborhood-level, SDoH such as park access and Area Deprivation Index (ADI) are associated with mortality in sepsis. Incorporation of these variables may assist in patient risk stratification, potentially improving public health initiatives or allowing them to be more targeted to at-risk populations.

## Introduction

Sepsis is a life-threatening medical emergency that occurs when the body’s response to an infection damages its own tissues and organs. Each year, approximately 1.7 million adults in the United States develop sepsis, and over 350,000 of them succumb to the condition.^1^ Socio-demographic factors, including economic status, race, and geographic location, may significantly influence sepsis outcomes.^2^

The World Health Organization defines “social determinants of health” (SDoH) as “the conditions in which people are born, grow, live, work, and age, along with the systems established to address illness.^3^” Although access to healthcare is well known to influence mortality, the impact of SDoH on sepsis outcomes is less well understood. SDoH encompasses community-level factors, like neighborhood socio-economic status and healthcare infrastructure, and individual-level variables such as education, family support, and access to transportation and healthcare. These variables are as vital to personalized medicine as the genetic or molecular profiles derived from ‘omics’ analyses. Current research highlights that SDoH are under-utilized in studies evaluating the long-term effects of sepsis, indicating a need for more transparent and inclusive data models to better support research and clinical applications.^4^

Given the burden of sepsis on morbidity, mortality and healthcare costs, gaining a clearer understanding of how SDoH interact with sepsis outcomes could inform the development of improved screening and management strategies to reduce adverse effects of the disease.^5,6^ Furthermore, the expanded use of electronic health records (EHR) and automated risk assessment tools has prompted renewed efforts to incorporate SDoH into predictive algorithms.^7,8^ These advancements have the potential to support clinicians in personalizing care and ultimately improving patient outcomes.

This study aims to address existing gaps in the literature regarding the relationship between SDoH and sepsis mortality rates among adults in the United States. Although the Surviving Sepsis Campaign provides comprehensive guidelines for sepsis management, it does not include recommendations for identifying patients at heightened risk of sepsis-related mortality based on social factors.^9^ By leveraging socioeconomic and patient data from the United States, our descriptive analysis seeks to provide healthcare professionals with deeper insights into patient risk, ultimately aiding in the identification of the most vulnerable populations and informing targeted care strategies.

## Methods

### Study Design and Setting

We used data from the TriNetX Diamond Network, a global health research network providing a de-identified dataset of electronic medical records (diagnoses, procedures, medications, laboratory values, genomic information) from (i) patients aged 18 years or older, (ii) who had been admitted to a hospital for medical care (as defined by hospital billing codes), and (iii) who had been diagnosed with sepsis by International Classification of Diseases v10 (ICD-10) codes (see Appendix). Location was assessed using the first three digits of ZIP codes available in the dataset. To preserve de-identification, the age of patients over 90 years of age was recorded as 90. Approximately 1% of patients lacked ZIP code information, and only 77.9% (3,436,375 patients) had valid ZIP codes.

The data was de-identified based on standard defined in Section §164.514(a) of the HIPAA Privacy Rule. The process by which datasets were de-identified is attested to through a formal determination by a qualified expert as defined in Section §164.514(b)(1) of the HIPAA Privacy Rule. Protected Health Information was made available to the investigators on December 12, 2022, and included records from 4,454,130 patients.

### Variables

We utilized socioeconomic data from the Centers for Disease Control and Prevention (CDC), specifically ZIP Code Tabulation Area (ZCTA)-level SDoH metrics, derived from the 5-year estimates of the American Community Survey (ACS).^10^ These measures included: (i) persons aged 65 years or older, (ii) households lacking broadband internet access, (iii) housing unit crowding, (iv) housing cost burden, (v) adults aged 25 years or older without a high school diploma, (vi) individuals living below 150% of the federal poverty level, (vii) persons of racial or ethnic minority status, (viii) single-parent households, and (ix) unemployment among people aged 16 years or older in the labor force. Supplementary Table 1 defines each of these features.

Additional data elements included income at the ZCTA level,^11^ park access at the county level,^12^ and the Area Deprivation Index (ADI) at the census tract level.^13^ ADI is a composite metric derived from 17 indicators, such as poverty, housing, education, and employment, that ranks neighborhoods within each state based on social disadvantage. Taken together, these variables provided a comprehensive indication of individual and neighborhood-level SDoH.

### Geographic Mapping

A key challenge in this study was integrating geographic data, as it was defined inconsistently across different datasets. ZIP codes, primarily designed for postal delivery, do not always align with census tract or county boundaries, which are the units commonly used for statistical and administrative purposes by government agencies. To address this misalignment, we used crosswalk tables,^14^ which are reference tools that map data between different geographic units, such as ZIP codes, census tracts, and counties. After crosswalk table mapping, all aggregated data were classified according to the first three digits of the ZIP code, ensuring consistency in all subsequent analyses.

### Statistical Analysis

Mortality rate was defined by the number of deceased patients divided the total number of patients within each ZIP code. The median mortality rate was then used to define areas having high or low mortality rates (HMR and LMR, respectively). To improve data quality, outliers were identified and excluded using a threshold of three times the interquartile range. The remaining data were standardized through scaling and centering.

We compared HMR and LMR groups using the Mann-Whitney U test, suitable for analyzing continuous variables. To investigate associations between outcomes and predictors, we began with univariate analyses for each predictor using binomial generalized linear models (GLMs). Predictors with a *P*-value less than 0.1 in the univariate analysis were then included in a multivariate binomial GLM to evaluate their combined effects. In the multivariate analysis, we calculated the Variance Inflation Factor (VIF) to identify multicollinearity among predictors, excluding those with a VIF greater than 5 to enhance model stability and interpretability.

Analyses were performed using Python 3.11 and the *statsmodels* package,^15^ with statistical significance set at *P* < 0.05.

## Results

### Comparison of Social Determinants of Health Between High- and Low-Mortality Neighborhoods

HMR neighborhoods exhibited significant differences in SDoH compared to LMR neighborhoods (Table 1). Median income in HMR neighborhoods was significantly lower ($61,754, IQR = $16,831) than in LMR neighborhoods ($71,051, IQR = $27,262, p < 0.001). Furthermore, the univariate GLM results (Table 2) show a negative coefficient for income (−0.69, 95% CI = [−0.86, −0.52], p < 0.001), confirming that lower income is strongly associated with higher sepsis mortality rates. Supplementary Figure 1 highlights the strength of this association, showing that income has one of the strongest negative correlations with mortality among the evaluated SDoH variables. This finding is consistent with existing evidence linking income disparities to access to healthcare, nutritious food, and stable housing.

**Table 1.** Comparison of Social Determinants of Health Between High and Low Mortality Rate neighborhoods. HMR = high mortality rate neighborhood; LMR = low mortality rate neighborhood.

| **Variable** | Missing values (%) | HMR, Median | HMR, IQR | LMR, Median | LMR, IQR | Median of all patients | *P*-Value |
| --- | --- | --- | --- | --- | --- | --- | --- |
| Crowding among housing units (%) | 1.0 | 0.8 | 1.1 | 1.8 | 2.1 | 1.3 | < 0.001 |
| Housing cost burden among households (%) | 1.0 | 20.0 | 5.2 | 23.8 | 8.9 | 21.3 | < 0.001 |
| No broadband internet subscription among households (%) | 1.0 | 18.0 | 6.6 | 12.9 | 8.0 | 15.7 | < 0.001 |
| No high school diploma among adults aged 25 years or older (%) | 0.1 | 9.6 | 6.9 | 8.4 | 5.9 | 9.0 | < 0.001 |
| Persons aged 65 years or older (%) | 0.0 | 19.0 | 3.3 | 16.6 | 5.4 | 18.0 | < 0.001 |
| Persons living below 150 % of the poverty level (%) | 0.9 | 21.4 | 10.2 | 19.6 | 9.5 | 20.3 | < 0.001 |
| Persons of racial or ethnic minority status (%) | 0.0 | 8.4 | 19.5 | 30.3 | 35.1 | 19.1 | < 0.001 |
| Single-parent households (%) | 1.0 | 4.3 | 2.0 | 4.8 | 2.5 | 4.5 | < 0.001 |
| Unemployment among people 16 years and older in the labor force (%) | 0.5 | 4.3 | 2.0 | 4.8 | 2.3 | 4.5 | < 0.001 |
| Mortality rate | - | 0.4 | 0.1 | 0.3 | 0.1 | 0.3 | < 0.001 |
| Income (US dollars) | 9.3 | 61,754 | 16,831 | 71,051 | 27,262 | 65,507 | < 0.001 |
| Park access (%) | 0.0 | 27.7 | 32.3 | 64.07 | 44.8 | 43.4 | < 0.001 |
| Area Deprivation Index (rank) | 0.0 | 73.0 | 20.4 | 50.0 | 37.6 | 63.9 | < 0.001 |

**Table 2.** Generalized Linear Model Regression Results for Features Affecting Sepsis Mortality Rate.

|  | **Coefficient** | ***P*-value** | **95% Confidence Interval** |
| --- | --- | --- | --- |
| **Univariate Features** |  |  |  |
| Median Income | -0.69 | <0.001 | [-0.86, -0.52] |
| Park Access | -1.13 | <0.001 | [-1.3, -0.95] |
| Crowding among Housing Units | -1.12 | <0.001 | [-1.33, -0.91] |
| Persons of Racial or Ethnic Minority Status | -0.91 | <0.001 | [-1.10, -0.72] |
| No Broadband Internet Subscription | 0.89 | <0.001 | [0.71, 1.07] |
| Housing Cost Burden | -0.79 | <0.001 | [-0.97, -0.61] |
| Unemployment (16+ in Labor Force) | -0.27 | <0.001 | [-0.42, -0.12] |
| Single-parent Households | -0.24 | 0.001 | [-0.39, -0.09] |
| No High School Diploma (25+ Years) | 0.34 | <0.001 | [0.18, 0.49] |
| Persons Below 150% Poverty Level | 0.38 | <0.001 | [0.23, 0.54] |
| Persons Aged 65+ Years | 0.32 | <0.001 | [0.17, 0.47] |
| Area Deprivation Index | 1.17 | <0.001 | [0.97, 1.36] |
| **Multivariate Features** |  |  |  |
| Park Access | -0.58 | <0.001 | [-0.86, -0.31] |
| Crowding among Housing Units | -1.0 | <0.001 | [-1.32, -0.70] |
| Persons of Racial or Ethnic Minority Status | -0.84 | <0.001 | [-1.19, -0.48] |
| No Broadband Internet Subscription | 0.55 | <0.001 | [0.244, 0.85] |
| Housing Cost Burden | 0.19 | 0.21 | [-0.11, 0.51] |
| Unemployment (16+ in Labor Force) | 0.13 | 0.30 | [-0.13, 0.39] |
| Single-parent Households | 0.19 | 0.20 | [-0.10, 0.49] |
| No High School Diploma (25+ Years) | 0.24 | 0.14 | [-0.08, 0.58] |
| Persons Aged 65+ Years | -0.44 | <0.001 | [-0.70, -0.19] |

HMR neighborhoods also demonstrated higher housing cost burdens and reduced park access compared to LMR neighborhoods (Table 1). Park access, a significant predictor, had a strong negative association with mortality in the univariate analysis (*coef* = −1.13, 95% CI = [−1.30, −0.95], p < 0.001) and remained significant, though attenuated, in the multivariate model (*coef* = −0.58, 95% CI = [−0.86, −0.31], p < 0.001, Table 2). From Supplementary Figure 1, park access was the most negatively correlated factor with mortality (r = −0.54), further illustrating its potential role in mitigating adverse outcomes. Figure 2 illustrates the overlap between high sepsis mortality rates and regions with low park access. The diminished effect in the multivariate model suggests park access may act as a proxy for broader socioeconomic and environmental resources rather than functioning independently.

**Figure 1.**
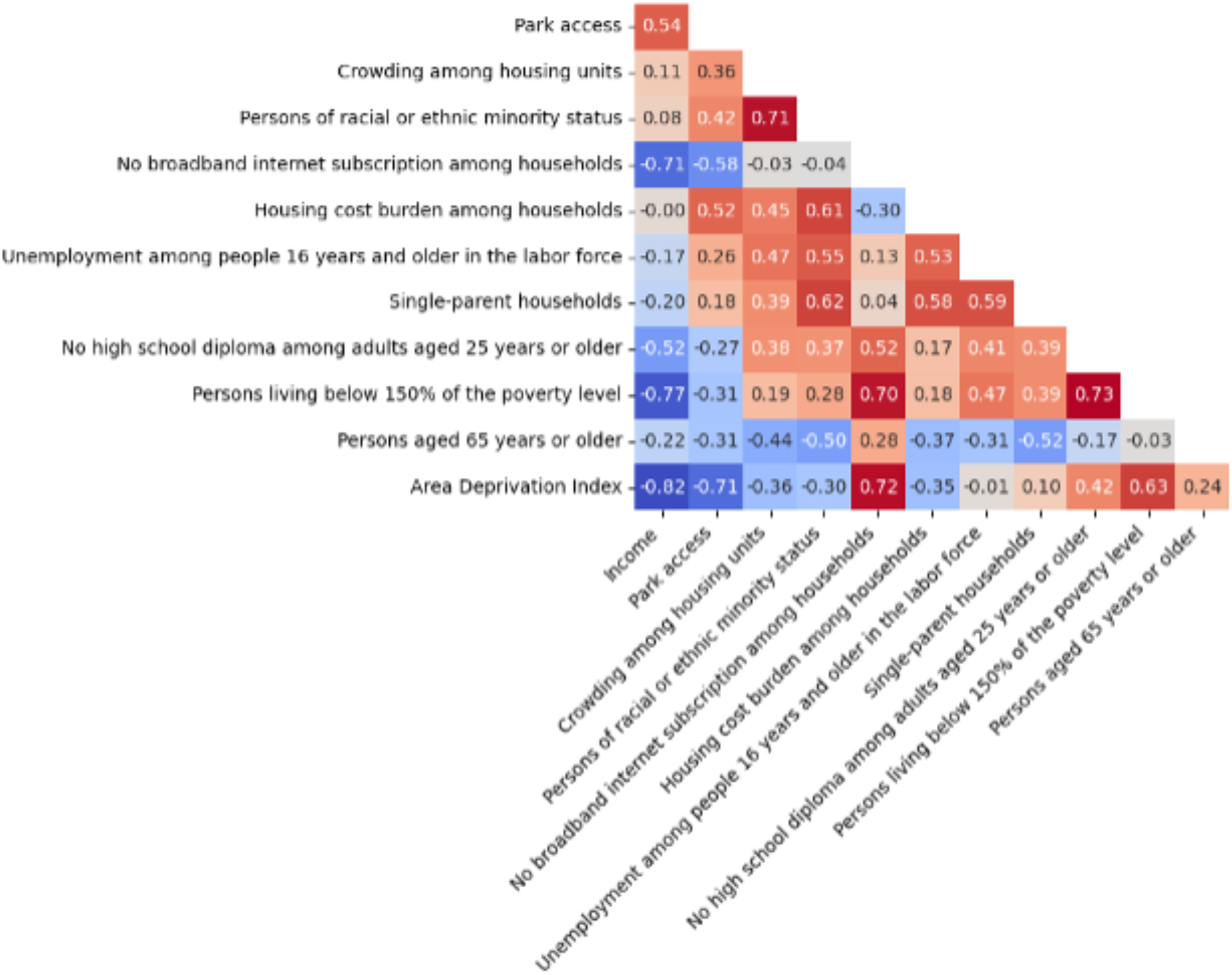
Heatmap of the correlation among Social Determinant of Health (SDoH) variables.

**Figure 2.**
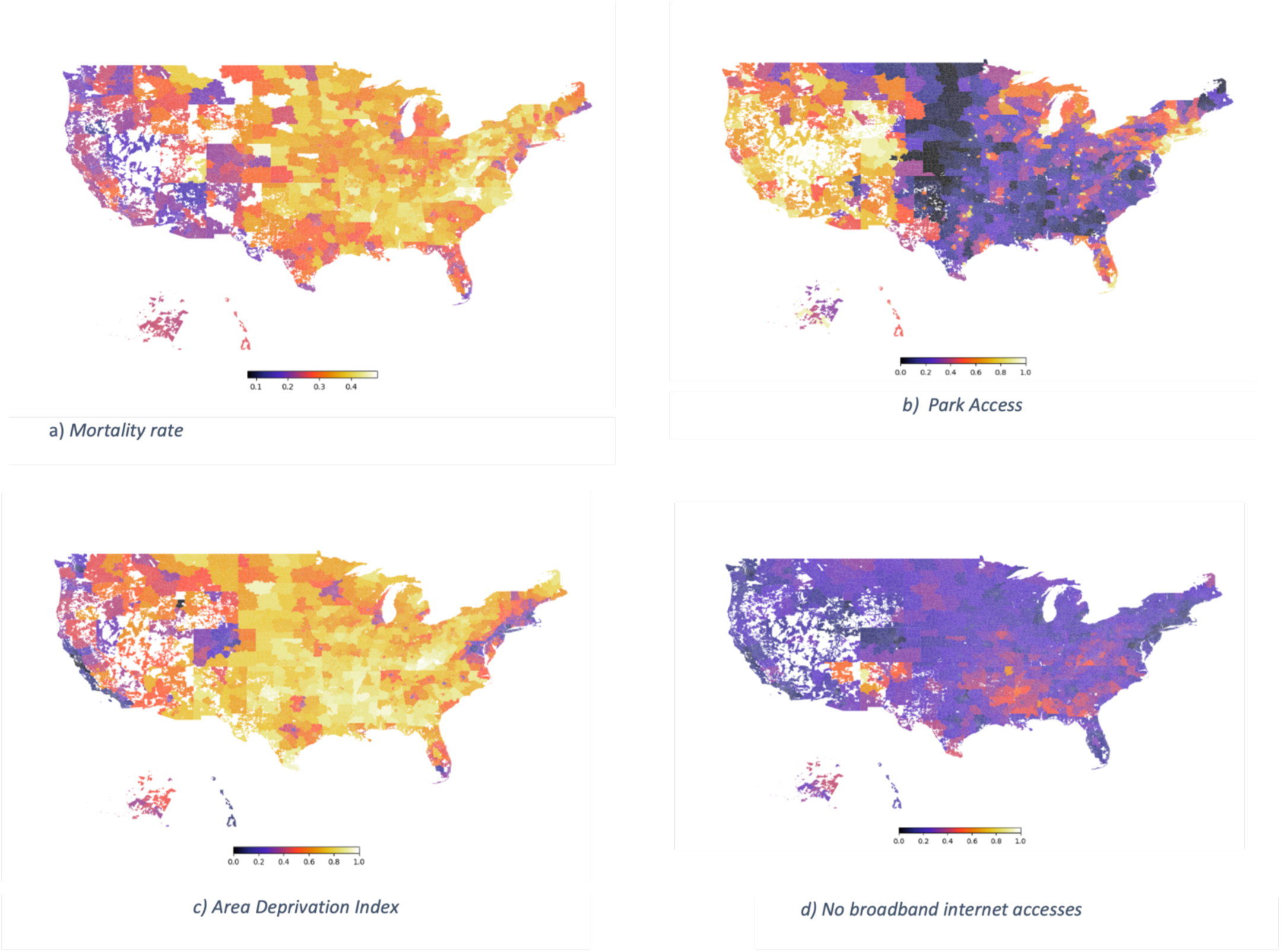
Map of the United States demonstrating sepsis mortality rate (a), together with predictors of sepsis mortality: (b) park access, (c) ADI, and lack of broadband internet access (d). Frequencies are normalized to a range between 0 and 1. Park access is based on the percentage of the population living within half a mile or one mile of a park.

Additional distinctions between HMR and LMR neighborhoods include higher ADI scores (median = 73.0 vs. 50.0, p < 0.001) and a larger proportion of households without broadband internet in HMR neighborhoods (Median = 18.0%, IQR = 6.6) compared to LMR neighborhoods (Median = 12.9%, IQR = 8.0, p < 0.001). Broadband availability had a significant positive association with mortality in both univariate (*coef* = 0.89, 95% CI = [0.71, 1.07], p < 0.001) and multivariate (*coef* = 0.55, 95% CI = [0.24, 0.85], p < 0.001) analyses, indicating that limited broadband access is a marker of increased sepsis mortality risk (Table 2).

HMR neighborhoods also had a higher percentage of older adults (Median = 19.0%, IQR = 3.3) compared to LMR neighborhoods (Median = 16.6%, IQR = 5.4, p < 0.001). However, the association between age and mortality shifted from positive in the univariate model (*coef* = 0.32, 95% CI = [0.17, 0.47], p < 0.001) to negative in the multivariate model (*coef* = −0.44, 95% CI = [−0.70, −0.19], p < 0.001), reflecting the influence of covariates like poverty and ADI.

In contrast, LMR neighborhoods were more racially and ethnically diverse, with a higher percentage of minority residents (Median = 30.3%, IQR = 35.1) compared to HMR neighborhoods (Median = 8.4%, IQR = 19.5, p < 0.001). Both univariate and multivariate models identified a negative association between racial/ethnic minority status and sepsis mortality (univariate *coef* = −0.91, 95% CI = [−1.10, −0.72], p < 0.001; multivariate *coef* = −0.84, 95% CI = [−1.19, −0.48], p < 0.001). This unexpected finding may reflect regional disparities, such as higher mortality in predominantly white rural areas facing unique challenges (e.g., hospital closures, limited transportation). Figure 1 illustrates interactions between racial diversity and other SDoH, suggesting a need for nuanced, context-specific interpretations.

Crowding was negatively associated with mortality in both univariate (*coef* = −1.12, 95% CI = [−1.33, −0.91], p < 0.001) and multivariate (*coef* = −1.00, 95% CI = [−1.32, −0.70], p < 0.001) models. These results suggest that increased residential density may serve as a protective factor in some settings, potentially due to improved access to shared resources or support networks in more crowded neighborhoods.

### Random Forest Analysis

To explore potential non-linear interactions, a Random Forest model was trained using 1,000 iterations, achieving mean accuracy of 74.65% (95% CI = [68.6%, 86.81%]) and an F1-score of 76.2% (95% CI = [69.68%, 81.97%]). Feature importance rankings (Figure 3) identified park access as the most predictive variable, aligning with its strong negative correlation with mortality (r = −0.54, Supplementary Figure 1). These findings reinforce the role of green spaces as critical components of public health strategies aimed at reducing mortality in disadvantaged communities.

**Figure 3.**
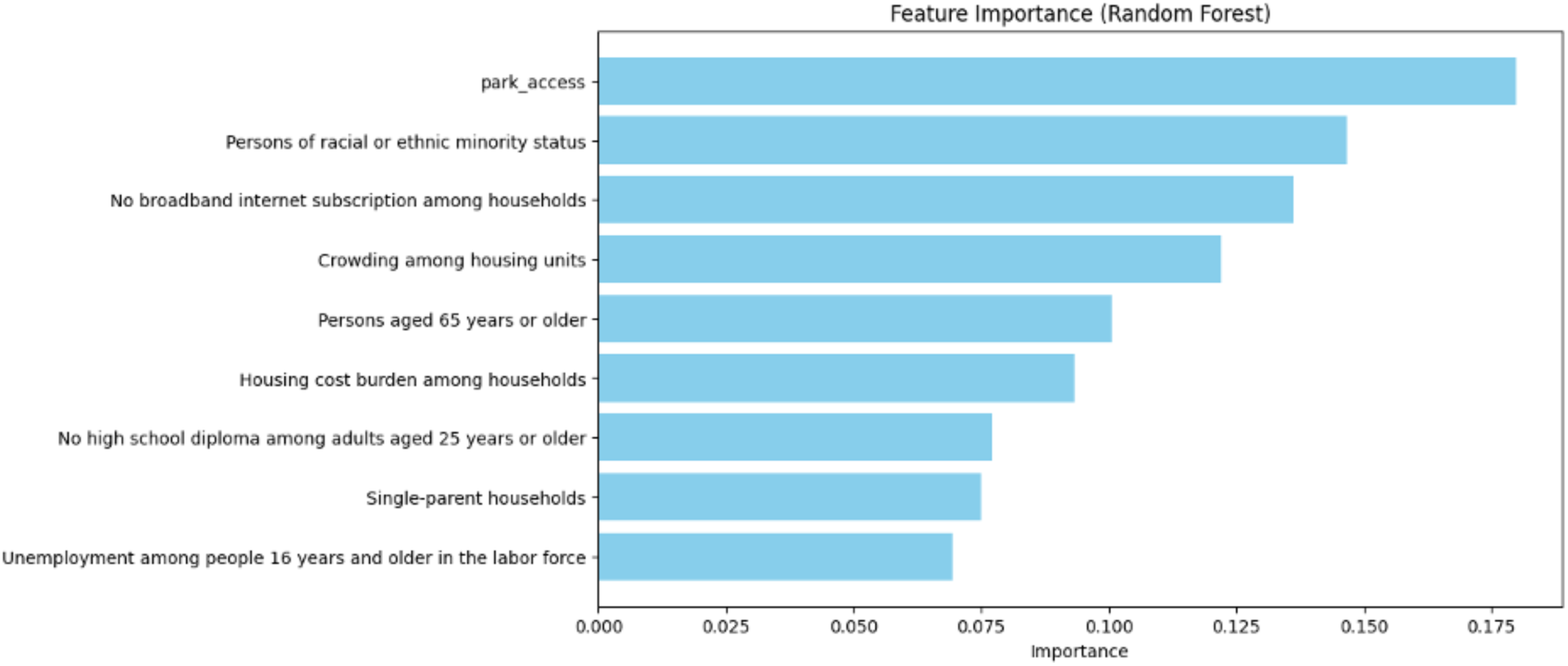
Relative feature importance in the prediction of high sepsis mortality rate. Analysis was performed using the Random Forest model.

## Discussion

Despite rapid advancements in machine learning, sepsis prognostic models have yet to achieve widespread clinical adoption. A primary barrier is limited model generalizability, often stemming from the exclusion of SDoH. Predictive models that fail to incorporate SDoH can yield biased or inaccurate outcome predictions. For example, studies have demonstrated that omitting SDoH from models leads to underestimation of hospitalizations across racial groups,^16^ whereas integrating SDoH improves in-hospital mortality predictions, particularly among Black patients.^17^ These findings underscore the necessity of incorporating SDoH into model frameworks to better reflect real-world patient populations and improve predictive accuracy.

The prominence of park access in both GLM and Random Forest analyses in this study highlights its potential as a proxy for broader neighborhood resources, such as infrastructure, community investments, and environmental quality. These results suggest that public health interventions targeting urban planning and environmental improvements—such as increasing access to green spaces—may play a pivotal role in reducing sepsis-related mortality, particularly in socioeconomically disadvantaged neighborhoods. Additionally, the associations we observed reinforce the critical role of SDoH in achieving equitable health outcomes. Patients from underprivileged communities are often underrepresented in clinical research due to limited data footprints, which restricts the development of models capable of addressing the needs of diverse populations. Incorporating SDoH into predictive models, particularly via EHR-embedded tools, has been proposed as a strategy to reduce disparities by accounting for patients’ broader social contexts.^19^

This study has several limitations. The use of aggregated data for de-identification purposes introduces the potential for ecological fallacy, where group-level associations may not necessarily apply to individual patients.^20,21^ However, the use of large datasets covering diverse geographic areas allows us to identify population-level trends and uncover social factors that may otherwise go undetected. Another limitation is residual confounding due to unmeasured variables. Although we excluded predictors with high multicollinearity (VIF > 5) to enhance model stability, attenuation of effects in the multivariate analyses (e.g., for park access and broadband availability) suggests shared variance among predictors that could not be fully accounted for.

Furthermore, the TriNetX network data is limited to participating healthcare organizations, which may exclude care provided outside these systems. Additionally, the dataset lacks detailed clinical characteristics and individual patient descriptors, which could affect the comprehensiveness and granularity of our findings. The cross-sectional nature of the study also restricts causal inference between SDoH and sepsis mortality. While the observed associations provide valuable insights, longitudinal studies are needed to establish temporal and causal relationships.

Future research should aim to incorporate a broader range of SDoH variables, including more granular data on transportation access, environmental exposures, and social cohesion. Longitudinal designs would enhance the ability to infer causality and evaluate the temporal relationship between SDoH and sepsis outcomes. Additionally, integrating SDoH directly into EHR systems at the patient level would address challenges related to combining community-level and patient-level data. Such integration could refine predictive models, increase their generalizability, and facilitate tailored interventions aimed at mitigating sepsis mortality. Leveraging advancements in natural language processing and geospatial analysis may also improve the precision of SDoH data extraction and application within clinical settings.^22,23^

In conclusion, this study highlights the importance of SDoH in sepsis outcomes and calls for their systematic integration into predictive modeling. Addressing disparities in sepsis care requires models that reflect the complexity of patients’ social environments, and these efforts could significantly improve outcomes for marginalized populations.

## Appendix

### Trinetx Query used for Study Inclusion

The following query parameters were used to generate the study cohort:

*Must Have: Age (≥ 18) AND (A41.9 Sepsis, unspecified organism OR A41 Other sepsis) AND (1013729 Critical Care Services OR 1013711 Emergency Department Services OR 1013648 Hospital Observation Services OR 1013659 Hospital Inpatient Services)*

### Ethics Approval

The study was conducted in accordance with institutional protocols established by the Penn State Human Subjects Protection Office (protocol# 17613, 5/10/2021).

## Data Availability

All data produced in the present work are contained in the manuscript

## Acknowledgements

This study was funded by the National Institute of General Medical Sciences, grant # R35GM150695 (ASB) and by the National Center for Advancing Translational Sciences (#UL1 TR002014). The content is solely the responsibility of the authors and does not necessarily represent the official views of the NIH.

