## Supplementary Material for "Social Determinants of Sepsis Mortality in the United States: A Retrospective, Epidemiologic Analysis"

### Supplementary Materials

**Supplementary Table 1**. Summary of ZIP Code Tabulation Area (ZCTA)-level social determinants of health (SDoH) measures.

| **Feature** | **Population** | **Numerator / Denominator** |
|---|---|---|
| Persons aged ≥ 65 years | Adults aged ≥ 65 years | Adults aged ≥ 65 years / All people for the same calendar year |
| No broadband internet subscription among households | All households | Households without broadband / All households for the same calendar year |
| Crowding among housing units | All occupied housing units | Occupied units with >1 occupant per room / All occupied units for the same calendar year |
| Housing cost burden among households | All households | Households with income <\$75,000 spending ≥ 30% on housing / All households for the same calendar year |
| No high school diploma among adults aged ≥ 25 years | Adults aged ≥ 25 years | Adults aged ≥ 25 years without a high school diploma / Adults aged ≥ 25 years for the same calendar year |
| Persons living below 150% of the poverty level | All people | People below 150% of poverty level / All people for the same calendar year |

| | | |
|---|---|---|
| Persons of racial or ethnic minority status | All people | People identifying as Hispanic or Latino (any race); Black and African American, Non-Hispanic; American Indian and Alaska Native, Non-Hispanic; Asian, Non-Hispanic; Native Hawaiian and Other Pacific Islander, Non-Hispanic; Other Races |
| Single-parent households | All households | Households with a single parent and children <18 / All households for the same calendar year |
| Unemployment among people ≥ 16 years in the labor force | People aged ≥ 16 years in civilian labor force | Unemployed people aged ≥ 16 years in civilian labor force / Population aged ≥ 16 years in civilian labor force |

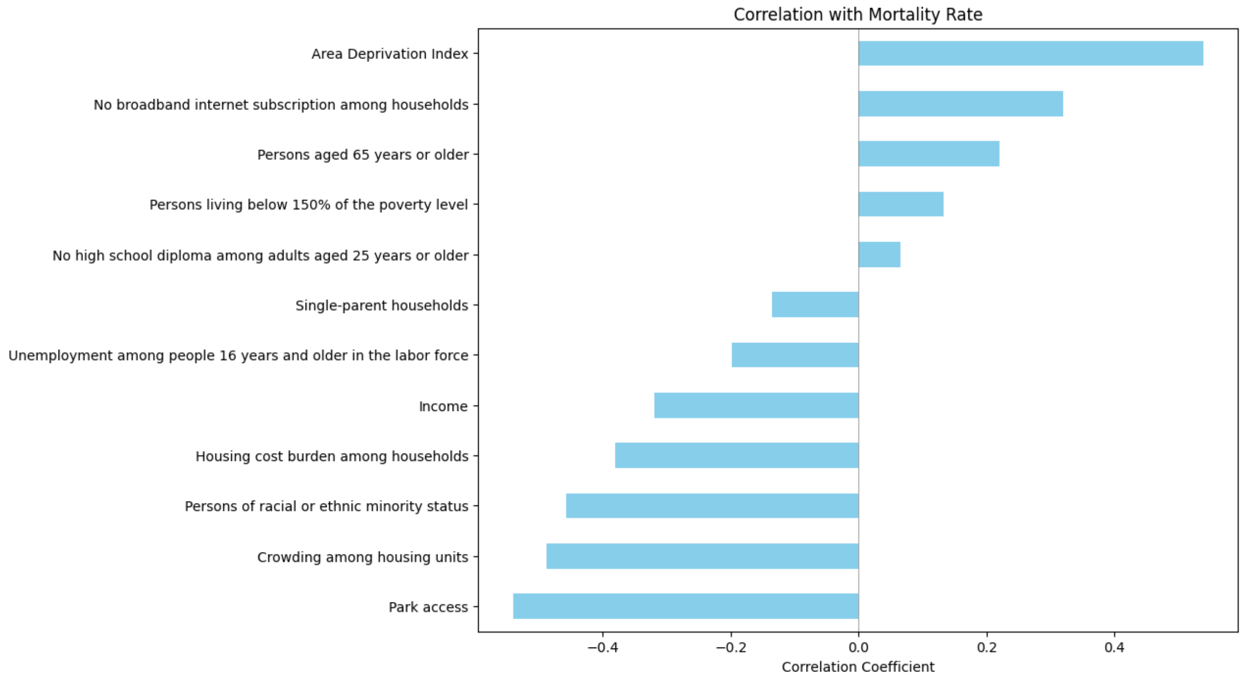


**Supplementary Figure 1.** Correlation between Social Determinants of Health (SDoH) and mortality rate. The top socioeconomic factor which has positive correlation with mortality rate is ADI (r = 0.53) where park access as shown here, the most negative association with the mortality rate (r= -0.54).
